# Assessing drug deposition efficacies, environmental impact and affordability for inhalers among chronic respiratory diseases: A systematic review

**DOI:** 10.1101/2024.06.05.24308532

**Authors:** Rahul Naresh Wasnik, Matthew Cope, Aaron Cowan, Smita Pakhale

**Author notes:** Corresponding author: Dr. Smith Pakhale MD, FRCPC, MSc (Epi & Biostat), Senior Scientist, Inflammation & Chronic Disease, Ottawa Hospital Research Institute, 501 Smyth Road, Ottawa, ON K1H 8L6, Canada., Ex. 79428.

## Abstract

**Introduction:** Chronic respiratory diseases such as asthma, COPD, and other pulmonary conditions impose a substantial global health burden, affecting millions of individuals worldwide. These conditions are characterized by persistent respiratory symptoms and reduced airflow, significantly impacting quality of life, and increasing healthcare needs. Treatment typically involves the use of inhaler devices to administer medications directly to the lungs, which decreases symptoms and improves outcomes. However, the efficacy of inhaler devices is influenced by various factors, including the type of device, patient adherence, and the correct device usage by patients.

In addition to clinical considerations, the environmental impact of inhaler devices, including their carbon footprint, as well as the cost implications for both healthcare systems and patients, are critical factors that require comprehensive evaluation. It is essential to develop sustainable and economically viable treatment strategies that address these considerations.

**Methods and Analysis:** We will conduct a systematic review aimed at providing a comprehensive understanding of the implications of inhaler use in treating asthma, COPD, and other chronic conditions requiring pulmonary drug delivery. Our focus will be on assessing efficiency, environmental sustainability, and cost-effectiveness. Studies lacking economic models or evaluations, without in-vivo deposition in the lungs, lacking assessment of the environmental impact of inhalers, not published in English, or falling into categories such as systematic reviews, letters, editorials, animal studies, or case studies will be excluded from this review.

The primary outcome of interest in this systematic review is the efficacy of medication deposition in the lungs of individuals with respiratory diseases when using different types of inhalers. The secondary outcome is to determine their economic costs and the tertiary outcome is to evaluate the overall environmental footprints of inhalers.

We will search for original research articles published until May 30, 2023, using databases such as MEDLINE (OVID), EMBASE (OVID), CENTRAL (OVID), the Canadian Drug and Health Technology Agency, and the US Food & Drug Administration. Our search method follows the PRISMA guidelines 2020. To determine study eligibility, a two-phase screening process will be conducted by three independent reviewers, with predefined outcomes extracted from eligible studies. The study will summarize findings through a narrative synthesis, using statistical analyses and sensitivity tests. In addition, funnel plots and Eggers test will be used for lung deposition analysis, while descriptive statistics will further compare costs and summarize environmental effects. We have ensured that a thorough risk of bias evaluation is part of this research approach, and it is carried out by three independent reviewers using a wide variety of tools according to the type of study.

**Ethics and Dissemination:** Ethics approval is not required for this study as it constitutes a protocol for a systematic review. The findings from this review will be disseminated via peer-reviewed publications and presented at conferences. Primary data will be available in the online repository on Open Science Framework, alongside a prior registration of this study.

**OSF Registration Number:** osf.io/xc5t6

https://doi.org/10.17605/OSF.IO/NT58B

**Strengths and limitations of this study:**

- The study will evaluate various aspects of inhaler use, including drug deposition efficiency, environmental sustainability, and cost-effectiveness.
- The findings aim to inform policy decisions and practice guidelines, focusing on the promotion of sustainable and economically viable healthcare solutions.
- The study seeks to enhance disease management, improve patient outcomes, and reduce healthcare costs.
- The study’s findings could be limited by the specific inhaler devices and patient populations examined, which may impact the conclusions’ generalizability.
- The fast pace of innovation in medical devices could mean that new inhaler technologies may emerge after the study’s completion that are not covered by the review but could have significant implications for patient care.

## INTRODUCTION

The global burden of asthma and chronic obstructive pulmonary disease (COPD) is on the rise in both high and low- and low-middle income countries. Recent estimates from the World Health Organization (WHO) indicate that around 235 million individuals suffer from asthma, while 65 million have moderate to severe COPD (Kibirige et al., 2017). Chronic respiratory diseases are *responsible* for approximately 4 million premature deaths annually, a figure that continues to grow, partly due to an aging population (Wisnivesky & de-Torres, 2019). Moreover, close to 2 billion people are exposed to harmful smoke from the combustion of biomass fuels in inadequately ventilated indoor environments, used for cooking or heating. Additionally, air pollution affects another billion, and a similar number are exposed to tobacco smoke, both directly and indirectly (Asher et al., 2021; Halpin et al., 2019). These factors significantly contribute to the global incidence of respiratory diseases, disproportionately affecting those of lower socio-economic status who face overcrowding, environmental hazards, and substandard living conditions *(Global Action Plan for the Prevention and Control of Noncommunicable Diseases 2013-2020, 2013)*.

The United Nations, recognizing the urgency of addressing non-communicable diseases (NCDs) including asthma and COPD, has prioritized their prevention and control in its 2030 agenda for sustainable development (Beaglehole et al., 2011; Meghji et al., 2021). A notable challenge in patient care has been the inefficient use of pressurized metered dose inhalers (pMDIs), where patients often struggle to synchronize device actuation with inhalation (Hirst et al., 2002). This challenge, coupled with the phasing out of chlorofluorocarbon propellants, has led to the development of dry powder inhalers (DPIs). DPIs are considered more efficient in delivering medication to the lungs than pMDIs and represent a promising advancement in pulmonary drug administration (Ball et al., 2002; Hirst et al., 2002; Leach et al., 1998; Newman SP; Hirst PH; Pitcairn GR, 2001). The propellant-free multidose inhaler device (SMI) reduces aerosol deposition in the mouth-throat region by eliminating the “ballistics effects” associated with pMDI’s. Medication stored in as a liquid in sealed systems such as prefilled syringes or cartridges within SMI’s. Upon Activation, mechanical energy is utilized to release the drug as gentle aerosol cloud (Newman SP; Hirst PH; Pitcairn GR, 2001; Sorino et al., 2020). This specific characteristic of SMI’s allow for a reduction in the dose while achieving therapeutic efficacy and improved safety where as pMDI’s are suitable for both maintenance and reliever with a short treatment time and high probability (Komalla et al., 2023). The DPIs are environmentally friendly than pMDI’s as they do not use propellants (Komalla et al., 2023; Woodcock et al., 2022).

A variety of inhaler devices are used for asthma and COPD treatment, and the wide breadth available impacts treatment decisions made by clinicians and patients. These decisions go on to affect both individual’s health and healthcare costs. Studies show differences in lung deposition percentages between devices, with Turbuhaler delivering approximately twice the drug dose to the lungs compared to others (Brocklebank et al., 2001). Dry powder inhalers are more effective than pressurized metered dose inhalers with a spacer in improving peak expiratory flow in COPD is limited to patients with severe symptoms, significant airflow restriction and frequent exacerbations in the past year. More data is needed to assess its benefits in patients with mild to moderate COPD who haven’t previously received treatment (Takahashi et al., 2023). While each device is promoted to have an advantage over its competitors, there is limited evidence to substantiate claims of superior efficacy for any specific device (Barry & O’Callaghan, 2003).

The economic burden of chronic respiratory diseases is substantial, encompassing both direct costs, such as those for devices and medication, and indirect costs, including those related to exacerbations and healthcare services (Håkansson et al., 2023; Usmani & Levy, 2023). This financial strain is further underscored by the WHO’s strategy for managing chronic respiratory diseases emphasizes enhancing healthcare through cost-effective interventions, improving care standards, expanding access, and ensuring the affordability of medications (*WHO Strategy for Prevention and Control of Chronic Respiratory Diseases*, 2002). In light of this, asthma, for instance, imposes an annual economic burden of approximately $81.9 billion in the United States alone, with a significant portion of this cost attributed to the high price of inhalers (McCarthy et al., 2022). In Canada, chronic lung diseases accounted for over 6% of the country’s annual healthcare expenses, totalling Canadian $12 billion in 2010 (Johnston et al., 2021; *The Impact of Lung Disease*, n.d.). Similarly, in the European Union, asthma and COPD represent a significant portion of the direct healthcare costs associated with respiratory diseases (Burney et al., 2015). Recent advancements in inhaler technology necessitate a re-evaluation of their role in patient care, particularly considering their environmental impact and economic implications (Usmani & Levy, 2023; Wilkinson & Woodcock, 2022).

Recent advancements in inhaler technology also necessitate a re-evaluation of their role in patient care, particularly considering their environmental impact. The carbon footprint of inhaler devices, especially those using hydrofluoroalkanes (HFAs) as propellants, has sparked concerns about their ecological consequences (Janson et al., 2020; Wilkinson & Woodcock, 2022). Globally, the release of HFCs from MDIs in 2014 was estimated to be equivalent to 13 million tonnes CO2, accounting for about 3% of the global warming potential-weighted CO2 emissions of HFCs (Montreal Protocol on Substances That Deplete the Ozone Layer, 2015). Transitioning from CFC to HFA propellants has the potential to significantly reduce their carbon footprint. Despite this, the overall contribution of pMDIs to greenhouse gas emissions is less than 0.1% (Pernigotti et al., 2021). Annually, the use of 630 million MDIs worldwide results in an estimated 13 million tonnes of CO2 equivalent emissions, highlighting the importance of developing low-GWP propellant devices and alternative inhalers with reduced carbon footprints (Kponee-Shovein et al., 2022). As the healthcare sector moves towards more sustainable practices, assessing the environmental impact of inhaler use is crucial for aligning with global sustainability goals (Usmani & Levy, 2023). The environmental impact of inhalers is significant with a single inhaler having a carbon footprint equivalent to a 20-mile car journey (Dipper et al., 2018). So, the majority of the inhalers are not recycled and are disposed of inappropriately through domestic waste (De Vos et al., 2020). The plastic waste from inhalers contributes to landfill and overtime, releases residuals HFC’s into the atmosphere (Dipper et al., 2018; Mikolasch & Stadler, 2020). Click or tap here to enter text.. This systematic review aims to address the need for improved inhaler devices by focusing on the efficiency of drug deposition into the lungs, the affordability of these devices, and the environmental impact of inhaler usage. Through a comprehensive examination of these aspects, the review seeks to contribute to the optimization of patient care within a sustainable and accessible framework. The objectives are to assess the impact of various inhaler devices on patient treatment outcomes, investigate the ecological consequences of their use, and analyze economic factors to identify barriers to affordable treatment. The findings of this review could inform global policy decisions by advocating for the development and promotion of inhaler devices that optimize drug deposition, minimize environmental impact, and improve affordability, thus advancing patient care with sustainable healthcare frameworks. Future directions may involve targeted interventions to address identified barriers and promote the widespread adoption of efficient, eco-friendly, and cost-effective technologies in clinical practice.

## OBJECTIVES

We will perform this systematic review to (a) assess the efficiency of drug deposition into the lungs of individuals by various inhaler devices in patients with chronic respiratory diseases, emphasizing the direct impact on patient treatment outcomes (b) investigate the environmental impact of different inhaler systems, including their contributions to greenhouse gas emissions and overall environmental footprint (c) analyze the financial costs associated with the use of inhalers by individuals with chronic diseases, considering both direct and indirect costs, and to compare the economic implications of different inhaler types and delivery systems.

## METHODS

We will follow the 2020 PRISMA guidelines for systematic review protocols to develop our search strategy, conduct our research, and present our findings (see online supplementary file 1) (Page et al., 2021). A reference copy of the completed checklist and search strategy will be included in the published article for convenient access.

### Study Registration

This study is registered a prior on Open Science Framework (osf.io/nt58b).

### Eligibility Criteria

The studies to be included in this review will consist of prospective studies, retrospective studies, and randomized controlled trials (RCTs). We will consider only original research articles published in English for inclusion in this analysis. Regarding our primary objective in evaluating the lung deposition of medication, we will include any studies that report primary outcomes on medication deposition in the lungs, as measured by gamma scintigraphy, single-photon emission computed tomography, positron emission tomography, bronchoalveolar lavage analysis or other methods. We will exclude studies that examine medication deposition by evaluating plasma, serum or urine levels as physiologic clearance of different individuals may confound the results of inhaler efficacy. Regarding our other objective of financial costs, we will include any studies that estimate the cost-effectiveness of inhaler-based medications, including medication expenses, healthcare visits, hospitalizations, emergency visits, costs associated with the devices, as well as indirect costs such as loss of productivity and out-of-pocket expenses. Regarding environmental impact, we will include all studies that examine quantitative measures of inhalers on the environment, such as emissions of greenhouse gasses, energy consumption during production, packaging materials, and overall environmental footprint.

### Participants

The participant demographics for studies exploring the efficiency of drug deposition or the financial costs associated with inhaler use will encompass individuals of any age diagnosed with chronic respiratory conditions, notably asthma and COPD, who rely on inhalers for their treatment. Studies assessing the environmental impact of inhalers will engage a broader participant base, inclusive of the general population affected by these devices. This not only pertains to the patients directly using inhalers but also includes the wider community that may experience the environmental repercussions stemming from inhaler use.

### Search Strategy

We will collaborate with a medical librarian (Risa Shorr, the Ottawa Hospital Research Institute) to develop a comprehensive search strategy with guidance from the PRISMA 2020 protocol checklist. Detailed searches will be conducted across multiple databases such as MEDLINE (OVID), EMBASE (OVID), CENTRAL (OVID), the Canadian Drug and Health Technology Agency, and the US Food & Drug Administration. While the search strategy planned to retrieve studies before May 30^th^, 2023, it will be updated closer to the final publication date of this systematic review to ensure any recent studies are included and to address any concerns regarding potentially missing recent literature. We will conduct detailed searches using appropriate keywords and will be supplemented as material 2. Searches will only consider publications that meet inclusion criteria and were published in English. The bibliographic management software Covidence will be employed to streamline the organization and screening of the retrieved articles. We will search thoroughly on several databases for full study texts. Studies for which we cannot find full texts will be excluded.

### Study Selection

After conducting pilot exercises using Covidence to resolve any discrepancies in the application of the inclusion and exclusion criteria, three independent reviewers working in pairs will systematically manage and evaluate the titles and abstracts of retrieved articles. Pilot tests will be run until a kappa-agreement value of 0.8 or higher is achieved between all reviewers. This preliminary phase aims to filter out studies that clearly do not meet the inclusion criteria. Studies without abstracts or with ambiguous titles will be provisionally advanced to full-text screening (second phase) for a more detailed evaluation unless both reviewers can determine that the study can be excluded at this stage. In the second phase, the same reviewers, working in pairs, will scrutinize the full texts of the shortlisted studies against the review’s inclusion and exclusion criteria. This thorough examination will be critical in determining the studies’ eligibility for the final analysis. Throughout both phases, any discrepancies between the reviewers will be flagged by Covidence for further discussion. In instances where consensus cannot be reached, a third reviewer will be consulted to mediate and resolve conflicts. .

#### Inhaler Lung Deposition

Studies are included if they investigate in vivo deposition in the lungs through various methodologies such as randomized controlled trials, prospective studies, or retrospective studies, and specifically involve inhalers. Studies are excluded if they do not examine in vivo lung deposition, are case studies, animal studies, systematic reviews, editorials, reviews, letters, are not published in English, or do not involve inhalers.

#### Inhaler Costs

To be included, studies must provide an economic evaluation or economic model related to inhalers and involve inhalers directly. Studies are excluded if they lack an economic evaluation or model of inhalers, are case studies, animal studies, systematic reviews, editorials, reviews, letters, do not involve inhalers, or are not published in English.

#### Inhaler Environmental Impact

Inclusion criteria require studies to assess the environmental impact of inhalers and to involve inhalers. Exclusion criteria cover studies that do not evaluate the environmental impact of inhalers, as well as case studies, animal studies, systematic reviews, editorials, reviews, letters, studies that do not involve inhalers, or those not published in English.

### Types of Interventions

#### 1. In vivo Lung deposition

Studies examining the efficiency of drug deposition in the lungs following the use of various inhalers including MDI or DPI’s. The intervention targets individuals of any age with chronic respiratory diseases necessitating the use of inhalers for drug delivery. The primary target is on the intervention of inhaler use (e.g.: MDI or DPIs) or different techniques for inhaler use (e.g.: spacer devices, breath-actuated inhalers) will also be considered to provide insights into comparative effectiveness and potential variations in drug deposition efficiency.

#### 2. Financial Costs

The economic models or evaluations estimate the cost-effectiveness of inhalers and inhaler medications. This will also assess the direct cost (medication expenses, healthcare visits, hospitalizations, emergency visits, costs related to inhaler devices and accessories) and indirect cost (loss of productivity, out-of-pocket expenses). This will compare the cost associated with various inhalers, as well as variations in different healthcare settings (e.g. primary care, specialty clinic, hospital settings). Additionally, comparisons between brand-name and generic inhalers will be explored to understand potential cost savings strategies.

#### 3. Environmental impact

Studies evaluating the environmental impact of inhalers encompass quantitative measures, such as emissions of greenhouse gas emissions, energy consumption during the production of inhalers, packaging materials. and overall environmental footprints associated with the lifecycles of different inhaler systems.

### Types of Outcomes

The efficiency of drug deposition will be evaluated by measuring how effectively medications are delivered into the lungs. Analyzing the financial costs of inhalers will encompass direct and indirect costs associated with inhaler use, including medication costs, healthcare visits, hospitalizations, lost productivity, and out-of-pocket expenses for patients. Lastly, assessing the environmental impact of inhalers will entail quantifying measures such as emissions of greenhouse gases per dose or the overall environmental footprint of different inhaler systems.

### Risk of Bias Assessment

To guarantee the validity and reliability of the study findings, a thorough risk of bias assessment will be carried out as part of this research. Three reviewers will evaluate the risk of bias independently by applying different risk of bias analysis tools based on the type of study.

Randomized controlled trials focused on inhaler deposition will employ Risk of Bias 2 (RoB 2) methodology, integrated within Covidence Revman. The Risk of Bias in Non-randomized Studies of Interventions (ROBINS-I) tool will be used for non-RCTs investigating inhaler deposition (J. Sterne et al., 2023). Confounding, selection of participants, classification of interventions, deviations from intended interventions, missing data, measurement of outcomes, and selection of reported results are all covered by ROBINS-I. With this tool, we will more accurately gauge the risk of bias in non-randomized studies and make sure that any potential sources of bias are thoroughly investigated and disclosed. The Philips technique (Philips et al., 2004)will be used for economic evaluations to determine the likelihood of bias in economic research. To assess the environmental impact of inhalers, the Guidelines for Life-Cycle Assessment: A “Code of Practice” will be used (Consoli et al., 1993).

The comprehensive methodology employed for evaluating the possibility of bias will enhance the overall strength and reliability of the research outcomes. In case of disagreement, a fourth author will be consulted.

### Data Analysis

Firstly, a narrative synthesis summarising the features and findings of separate objectives in the study will be presented in the text and in tables. Statistical analyses of lung deposition, economic impact, and environmental impact will include heterogeneity assessment, where subgroup analyses based on appropriate variables may be carried out to investigate potential sources of heterogeneity.

Statistical measures such as the I^2^ statistic will be utilized to assess heterogeneity among studies. In addition, sensitivity analyses will be conducted to evaluate the reliability of the deposition, environmental, and economic conclusions. Sensitivity analysis will also investigate the effects on the overall results of removing studies that have a high risk of bias or substantial methodological constraints.

To determine the significance of variations in results between groups, statistical tests (such as chi-squared tests and t-tests) will be employed. Cochrane’s RevMan and R programming will be used for all statistical tests. Analyses specific to lung deposition studies will include funnel plots to visually evaluate publication bias, and Egger’s test for publication bias if appropriate.

To compare the direct and indirect costs related to various inhaler types, descriptive statistics will be employed. Where appropriate, cost-effectiveness ratios will be computed. Finally, descriptive statistics will be used to summarise environmental effect data.

## DISCUSSION

To the best of our knowledge and understanding, this is the first systematic review to evaluate the need for improved inhaler devices with emphasis on drug deposition efficiency, environmental impact, and economic impact. We anticipate variations in study design among the publications included in our review. Nevertheless, we will accommodate this variability in our data analysis to ensure comprehensive and meaningful interpretations of our analyses.

The findings from this review will improve our understanding of the environmental implications, drug deposition, and economic constraints relating to different types of inhalers.

## Data Availability

All data produced in the present work are contained in the manuscript

https://doi.org/10.17605/OSF.IO/NT58B

## Funding

This research received no specific grant from any funding agency in the public, commercial or not-for-profit sectors

## Competing interests

The authors declare that they have no competing interests.

## Patient and Public involvement

As this is a protocol for a systematic review, the findings will be derived exclusively from previously published data. Patients and the public will not be involved in the design, conduct, report, or dissemination plans of this research.

## Patient Consent for Publication

Not applicable

## Author Contributions

RNW –writing the preliminary draft, Contributed to Risk of bias tools identification, preparation of-final version of manuscript

MC – editing and methods formatting, contributed to writing of risk of bias tools and data analysis section.

AC - editing and supervising the Manuscript, Contributed to Risk of bias tools identification

SP-Design, preparing, conceptualization, contributed to critically revised the final version of Manuscript

All authors read, edited, and approved the final version of the Manuscript

## Acknowledgments

Special thanks to Ryan Chow (Research Fellow, Ottawa Hospital Research Institute) for carrying out a final review of this manuscript and making suggestions on how to improve its quality, and Risa Shorr for creating the search strategy performing the literature searches.

## Supplementary files

Search Strategy:

### Inhalers - Lung Deposition

Ovid MEDLINE(R) ALL < 1946 to October 10, 2023 >;

1. exp &quot;Nebulizers and Vaporizers&quot;/ 12860
2. Administration, Inhalation/ and (Drug Delivery Systems/ or Equipment Design/ or exp Respiratory System Agents/ or particle size/) 17352
3. ((meter* dose or dry powder or soft mist*) adj3 inhal*).tw,kf. 6503
4. (nebuli?er* adj3 (design* or drug delivery or device*)).tw,kf. 249
5. (mdi adj3 (design* or drug delivery or device*)).tw,kf. 157
6. or/1-5 27603
7. ((drug or lung or pulmonary or aerosol* or central or peripheral* or oropharyng*) adj5 (deposit* or disposition)).tw,kf. 14043
8. inspiratory flow.tw,kf. or deposit*.ti. 46994
9. deposition*.ab. /freq=2 49415
10. 10 7 or 8 or 9 90595
11. 11 6 and 10 2526
12. exp animals/ not humans/ 5161760
13. 11 not 12 2293
14. limit 13 to english language 2199

### Embase Classic + Embase < 1947 to 2023 October 10>;

1. inhalational drug administration/ 51227
2. drug delivery system/ 172790
3. equipment design/ or pump design/ 94347
4. exp *respiratory tract agent/ or respiratory tract agent/ad 477418
5. *particle size/ 14795
6. 2 or 3 or 4 or 5 751950
7. 1 and 6 15169
8. exp nebulizer/ or respiratory therapeutic device/ 15322
9. inhaler/ or inhalation spacer/ or exp metered dose inhaler/ or exp powder inhaler/ 19383
10. exp *respiratory tract agent/ih, na [Inhalational Drug Administration, Intranasal Drug Administration] 10333
11. ((meter* dose or dry powder or soft mist*) adj3 inhal*).tw. 9654
12. (nebuli?er* adj3 (design* or drug delivery or device*)).tw. 465
13. (mdi adj3 (design* or drug delivery or device*)).tw. 259
14. or/7-13 55507
15. drug disposition/ 12163
16. ((drug or lung or pulmonary or aerosol* or central or peripheral* or oropharyng*) adj5 deposit*).tw. 14014
17. inspiratory flow.tw. or deposit*.ti. 53115
18. peak inspiratory flow/ 1620
19. deposition*.ab. /freq=2 64425
20. 15 or 16 or 17 or 18 or 19 118629
21. 14 and 20 3728
22. (exp animals/ or animal experiment/ or nonhumans/) not exp humans/ 6273337
23. 21 not 22 3469
24. limit 23 to english language 3269

### Inhalers – Environmental Impact

Ovid MEDLINE(R) ALL <1946 to October 10, 2023>;

1. exp &quot;Nebulizers and Vaporizers&quot;/ 12860
2. Administration, Inhalation/ and (Drug Delivery Systems/ or Equipment Design/ or exp Respiratory System Agents/ or particle size/) 17352
3. ((meter* dose or dry powder or soft mist*) adj3 inhal*).tw,kf. 6503
4. (nebuli?er* adj3 (design* or drug delivery or device*)).tw,kf. 249
5. (mdi adj3 (design* or drug delivery or device*)).tw,kf. 157
6. or/1-5 27603
7. environment/ or carbon footprint/ 68992
8. Greenhouse Effect/ 6170
9. Greenhouse Gases/ 2476
10. sustainability.tw,kf. 45883
11. (environment* adj2 (impact or friendly)).tw,kf. 39830
12. eco friendly.tw,kf. 12970
13. (carbon footprint* or greenhouse gas*).tw,kf. 16852
14. climate change/ 26918
15. climate change.tw,kf. 61577
16. environment*.ti. 205747
17. or/7-16 401423
18. 6 and 17 186
19. exp animals/ not humans/ 5161760
20. 18 not 19 181
21. limit 20 to english language 175

### Embase Classic + Embase < 1947 to 2023 October 10 >;

1. inhalational drug administration/ 51227
2. drug delivery system/ 172790
3. equipment design/ or pump design/ 94347
4. exp *respiratory tract agent/ or respiratory tract agent/ad 477418
5. *particle size/ 14795
6. 2 or 3 or 4 or 5 751950
7. 1 and 6 15169
8. exp nebulizer/ or respiratory therapeutic device/ 15322
9. inhaler/ or inhalation spacer/ or exp metered dose inhaler/ or exp powder inhaler/ 19383
10. exp *respiratory tract agent/ih, na [Inhalational Drug Administration, Intranasal Drug Administration] 10333
11. ((meter* dose or dry powder or soft mist*) adj3 inhal*).tw. 9654
12. (nebuli?er* adj3 (design* or drug delivery or device*)).tw. 465
13. (mdi adj3 (design* or drug delivery or device*)).tw. 259
14. or/7-13 55507
15. environmental impact/ or greenhouse effect/ 50367
16. greenhouse gas/ 8726
17. environmental sustainability/ 7871
18. (environment* adj2 (impact or friendly)).tw. 41950
19. sustainability.tw. 46869
20. eco friendly.tw. 13053
21. (carbon footprint* or greenhouse gas*).tw. 16927
22. climate change/ 59532
23. climate change.tw. 52011
24. environment*.ti. 244184
25. environment/ 125418
26. exp greenhouse gas emission/ 17608
27. or/15-26 540183
28. 14 and 27 690
29. (exp animals/ or animal experiment/ or nonhumans/) not exp humans/ 6273337
30. 28 not 29 671
31. limit 30 to english language 649

### Inhalers – Costs

Ovid MEDLINE(R) ALL < 1946 to October 10, 2023, >;

1. exp &quot;Nebulizers and Vaporizers&quot;/ 12860
2. Administration, Inhalation/ and (Drug Delivery Systems/ or Equipment Design/ or exp Respiratory System Agents/ or particle size/) 17352
3. ((meter* dose or dry powder or soft mist*) adj3 inhal*).tw,kf. 6503
4. (nebuli?er* adj3 (design* or drug delivery or device*)).tw,kf. 249
5. (mdi adj3 (design* or drug delivery or device*)).tw,kf. 157
6. or/1-5 27603
7. exp &quot;Costs and Cost Analysis&quot;/ 266638
8. (costs or cost effective*).tw,kf. or (&quot;Financial&quot; or &quot;Economic&quot; or &quot;Cost*&quot; or &quot;Affordability&quot; or &quot;Pricing&quot; or &quot;Expenditure&quot;).ti. 512814
9. 7 or 8 633989
10. 10 6 and 9 918
11. respiratory tract diseases/ or lung diseases/ or exp lung diseases, obstructive/ 326152
12. (lung disease* or lung health or pulmonary disease* or respiratory disease* or asthma* or copd or chronic obstructive pulmonary or respiratory tract disease*).tw,kf. 365284
13. 11 or 12 485467
14. 10 and 13 661
15. exp animals/ not humans/ 5161760
16. 14 not 15 658
17. limit 16 to english language 615

### Embase Classic + Embase < 1947 to 2023 October 10 >;

1. inhalational drug administration/ 51227
2. drug delivery system/ 172790
3. equipment design/ or pump design/ 94347
4. exp *respiratory tract agent/ or respiratory tract agent/ad 477418
5. *particle size/ 14795
6. 2 or 3 or 4 or 5 751950
7. 1 and 6 15169
8. exp nebulizer/ or respiratory therapeutic device/ 15322
9. inhaler/ or inhalation spacer/ or exp metered dose inhaler/ or exp powder inhaler/ 19383
10. exp *respiratory tract agent/ih, na [Inhalational Drug Administration, Intranasal Drug Administration] 10333
11. ((meter* dose or dry powder or soft mist*) adj3 inhal*).tw. 9654
12. (nebuli?er* adj3 (design* or drug delivery or device*)).tw. 465
13. (mdi adj3 (design* or drug delivery or device*)).tw. 259
14. or/7-13 55507
15. (cost or costs).tw. or (&quot;Financial&quot; or &quot;Economic&quot; or &quot;Cost*&quot; or &quot;Affordability&quot; or &quot;Pricing&quot; or &quot;Expenditure&quot;).ti. 1041677
16. *&quot;drug cost&quot;/ 11882
17. 15 or 16 1045999
18. 14 and 17 2343
19. asthma/ or obstructive lung disease/ 290214
20. chronic obstructive lung disease/ 177933
21. respiratory tract disease/ 80172
22. (lung disease* or lung health or pulmonary disease* or respiratory disease* or asthma* or copd or chronic obstructive pulmonary or respiratory tract disease*).tw. 557749
23. or/19-22 711635
24. 18 and 23 1543
25. (exp animals/ or animal experiment/ or nonhumans/) not exp humans/ 6273337
26. 24 not 25 1527
27. limit 26 to english language 1458

## Notes

### Competing Interest Statement

The authors have declared no competing interest.

### Funding Statement

This study did not receive any funding

